# SARS-CoV-2 in human milk is inactivated by Holder pasteurization but not cold storage

**DOI:** 10.1101/2020.06.18.20134395

**Authors:** Gregory J. Walker, Vanessa Clifford, Nidhi Bansal, Alberto Ospina Stella, Stuart Turville, Sacha Stelzer-Braid, Laura D Klein, William Rawlinson

## Abstract

As the COVID-19 pandemic evolves, human milk banks worldwide continue to provide donor human milk to vulnerable infants who lack access to mother’s own milk. Under these circumstances, ensuring the safety of donor human milk is paramount, as the risk of vertical transmission of SARS-CoV-2 is not well understood. Here, we investigate the inactivation of SARS-CoV-2 in human milk by pasteurisation, and the stability of SARS-CoV-2 in human milk under cold storage (freezing or refrigeration). Following heating to 63°C or 56°C for 30 minutes, SARS-CoV-2 replication competent (i.e. live) virus was undetected in both human milk and the control medium. Cold storage of SARS-CoV-2 in human milk (either at 4°C or - 30°C) did not significantly impact infectious viral load over a 48 hour period. Our findings demonstrate that SARS-CoV-2 can be effectively inactivated by Holder pasteurisation, and confirm that existing milk bank processes will effectively mitigate the risk of transmission of SARS-COV-2 to vulnerable infants through pasteurised donor human milk.

## INTRODUCTION

As the COVID-19 pandemic evolves, human milk banks worldwide continue to provide donor human milk to vulnerable infants who lack access to mother’s own milk.^1^ Under these circumstances, ensuring the safety of donor human milk is paramount, as the risk of vertical transmission of SARS-CoV-2 is not well understood.^2^ Although case series have reported that SARS-CoV-2 was not detected in breast milk,^3^ there have been recent case reports^4,5^ of SARS-CoV-2 detected in breast milk from women with symptomatic COVID-19 infection. Expressed milk may theoretically become contaminated from maternal respiratory secretions or via skin, though no evidence suggests that breast milk is a means of transmission of SARS-CoV-2 to infants. Consistent with available data, international guidelines recommend mothers with COVID-19 continue to provide breast milk for their babies as the benefits outweigh risks of virus transmission.^2^ Although low, the potential risk of SARS-CoV-2 transmission through milk is concerning for milk banks globally.^1^ Here, we investigate the inactivation of SARS-CoV-2 in human milk by pasteurisation, and the stability of SARS-CoV-2 in human milk under cold storage (freezing or refrigeration).

## METHODS

Frozen (14 weeks) and freshly expressed (<12 hours) human milk was obtained from healthy donors to Australian Red Cross Lifeblood Milk.

For the assessment of viral inactivation by pasteurisation, SARS-CoV-2 10^5^ TCID50/mL was diluted 1:10 in previously frozen human milk or non-supplemented Minimum Essential Medium (MEM, Gibco™, Life Technologies, CA) (control). Triplicates of each sample were layered onto uninfected cells after being subjected to Holder pasteurisation (63°C for 30 minutes), under-pasteurisation (56°C for 30 minutes) or were unpasteurised. Endpoint titration of samples was performed on Vero cells in sextuplicate, and the 50% tissue culture infectious dose (TCID50) determined at 72 hours post-infection using standard methods.^6^

Freshly expressed milk from two donors and MEM (control) was similarly inoculated with SARS-CoV-2 to assess virus stability under cold storage conditions. Infectious titres of samples were measured at the time of inoculation, and after 48 hours of storage at 4°C or -30°C. This project was approved by the Australian Red Cross Lifeblood HREC (Clifford 27042020).

## RESULTS

Following heating to 63°C or 56°C for 30 minutes, SARS-CoV-2 replication competent (i.e. live) virus was undetected in both milk and MEM (Fig. 1A). After 48 hours there was no reduction in SARS-CoV-2 infectious titre in milk samples kept at 4°C, and a 0.41 log-unit reduction in samples frozen at -30°C (Fig. 1B). In the control media (MEM), there was a 0.02 and 0.25 log-unit reduction in infectious titres of samples stored at 4°C and -30°C respectively. Whilst freezing of milk and MEM samples resulted in a slight reduction in infectious titre, viable virus was still recovered after 48 hours of storage.

**Figure 1.**
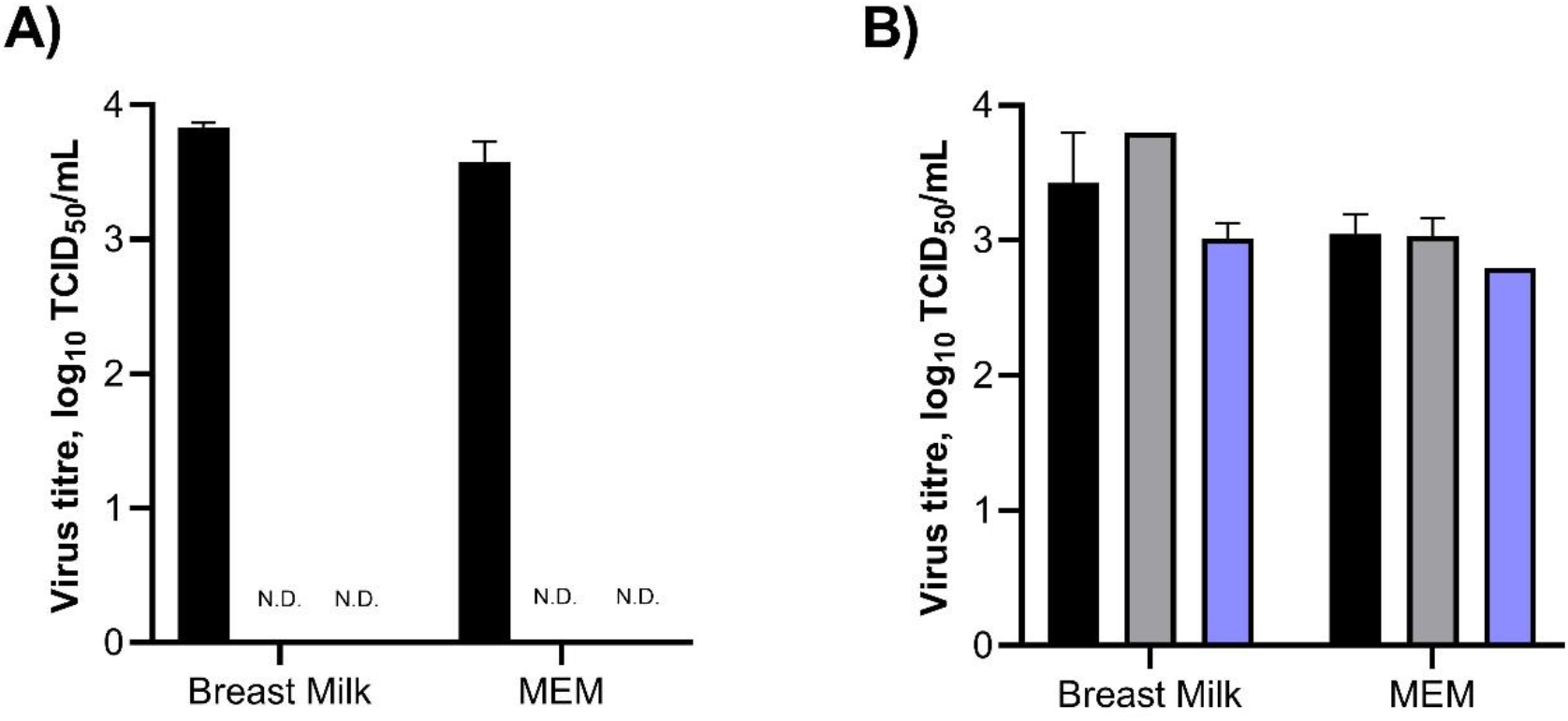
Viability of SARS-CoV-2 in breast milk. (A) Inactivation by pasteurisation. Previously frozen human breast milk or MEM was inoculated with SARS-CoV-2. Infectious titres were determined in triplicates of each sample that were subjected to Holder (63°C) pasteurisation (N.D.), under-pasteurisation (56°C) (N.D), or were unpasteurised (black). (B) Stability under cold storage conditions. Freshly expressed breast milk (n=2) or MEM (n=3) was inoculated with SARS-CoV-2. Infectious titres of samples were determined at 0 hours post-inoculation (black), and after 48 hours of storage at either +4°C (grey) or -30°C (blue). Infectious titres for all experiments were determined using endpoint titration on Vero cells in sextuplicate. Error bars indicate the standard error of the mean. *SARS-CoV-2, severe acute respiratory syndrome coronavirus 2; MEM, Minimum Essential Medium; N*.*D*., *not detected; TCID50, 50% tissue culture infectious dose*.

## DISCUSSION

Milk banks employ many risk mitigation strategies to ensure the safety of donor milk, with donor selection criteria, serological and/or viral nucleic acid screening, validated frozen transport methods, microbial testing, and pasteurisation.^7^ For SARS-CoV-2, this includes specific questions to screen for symptoms of COVID-19, deferral of donors with COVID-19 or close contact with COVID-19, and pasteurisation of donor milk. Holder pasteurisation (62.5°C for 30 minutes) is the most common pasteurisation method among milk banks worldwide, and has been shown to inactivate most enveloped viruses, including MERS-CoV^8^ and SARS-CoV-1.^9^ Our findings are consistent with a recent study that reported SARS-CoV-2 is inactivated by heat treatment,^10^ although inactivation under Holder pasteurisation has not yet been investigated in human milk. Our findings demonstrate that SARS-CoV-2 can be effectively inactivated by Holder pasteurisation, and confirm that existing milk bank processes will effectively mitigate the risk of transmission of SARS-COV-2 to vulnerable infants through pasteurised donor human milk.

This is the first study to assess the stability of SARS-CoV-2 in human milk. We show that cold storage of SARS-CoV-2 in human milk (either by refrigeration or freezing) does not significantly impact infectious viral load over a 48 hour period. Results of the refrigerated samples are consistent with a recent study not involving human milk that found SARS-CoV-2 to be highly stable at 4 ° C in the environment.^10^ Whilst it is yet to be determined whether SARS-CoV-2 detected in breast milk is infectious, these findings may assist in the development of guidelines around expressing and storing milk from COVID-19 infected mothers.

## Abbreviations

SARS-CoV-2: severe acute respiratory syndrome coronavirus 2
COVID-19: coronavirus disease 2019
MEM: minimum essential medium
TCID_50_: 50% tissue culture infectious dose

## Data Availability

The data that support the findings of this study are available from the corresponding author (WR), upon reasonable request.

## Acknowledgements

The authors thank Christine Sulfaro, Nicole Stevens, Lifeblood Milk donors, and members of the Serology and Virology Division (SAViD) Area Diagnostic Virology Laboratory and Kirby Institute UNSW for their assistance with this study.

## Notes

**Conflict of Interest Disclosures:** The authors have no conflicts of interest to disclose.

**Funding/support:** No funding was secured for this study.

### Competing Interest Statement

The authors have declared no competing interest.

### Funding Statement

No funding was secured for this study.

### Author Declarations

This project was approved by the Australian Red Cross Lifeblood Human Research Ethics Committee (Clifford 27042020).

